# Dysbiosis of Gut microbiota in patients with Large-Artery Atherosclerotic Stroke: a pilot study

**DOI:** 10.1101/2023.03.21.23287557

**Authors:** Chatpol Samuthpongtorn, Abhinbhen W. Saraya, Yutthana Joyjinda, Apaporn Rodpan, Nijasri C. Suwanwela

## Abstract

**Introduction:** Increasing data demonstrate an association between gut microbiome in brain diseases via the gut-brain axis. However, few studies have evaluated the association between gut microbiome and large-artery atherosclerotic ischemic stroke patients.

**Method:** A cross-sectional pilot study was conducted among 15 patients with large-artery atherosclerotic ischemic stroke and 15 asymptomatic persons. Large-artery atherosclerotic stroke were diagnosed using TOAST classification. The control group was selected based on age- and sex-match with the patient group. Participants provided a stool sample profiled by 16S-ribosomal gene-specific methods Next-generation sequencing. The Mann-Whitney U test was used to compare the differences in gut microbiota profile between stroke and control groups. Alpha (Shannon diversity index) and beta diversity (Bray-Curtis dissimilarity) were used to evaluate gut microbial diversity. Generalized linear mixed effects models were used to relate gut microbial genus and stroke which were adjusted for age, BMI, underlying disease (diabetes, hypertension and dyslipidemia), and alcohol use.

**Result:** The average age of stroke patients was 61.1±7.1 and 59.2±8.2 in the control group. Beta-diversity (Bray-Curtis dissimilarity) of the gut microbiome was statistically significant in order, family and genus level (P-value=0.017, 0.011 and 0.003, respectively) between stroke and control groups; however, there was no statistically significant difference in alpha-diversity (Shannon diversity index; P-value=0.852). The relative abundance of *Class Bacteriodia* increased in stroke group. Using generalized linear mix effect model, we found 6 genera was significantly associated with stroke after multivariate adjustment. *Ruminococcus spp*.(P-value=0.017), *Streptococcus spp*.(P-value=0.019), *Actinomyces spp*.(P-value=0.02) and *Dorea spp*.(P-value=0.021) showed positive association while *Bifidobacterium spp*.(P-value=0.04) and *Faecalibacterium spp*.(P-value=0.041) showed negatively association with stroke.

**Conclusion:** Patients with large-artery atherosclerotic stroke had a decreased microbiome beta-diversity and certain gut microbiota genera may be related to large-artery atherosclerotic stroke. Future implications of this study could include the development of targeted interventions to modulate gut microbiota in order to improve outcomes for patients with large-artery atherosclerotic stroke.

## Introduction

Stroke is the leading cause of disability, morbidity, disability adjusted life years (DALYs) lost and mortality for both women and men in Thailand and worldwide. In 2014, the prevalence of stroke among adults 45 years and older was estimated at 1.88%. The average age at which a stroke occurs was 65, which approximately 50,000 people die annually from a stroke ^1^. There are two most common types of stroke; ischemic stroke, which accounts for approximately 80% and hemorrhagic stroke, accounting for the remaining 20% of stroke cases in Thailand^2^. In 2010, DALYs lost from hemorrhagic stroke and ischemic stroke were around 62.8 million and 39.4 million, respectively^3^. In 2013, cerebrovascular disease(CVD) was the leading cause of DALYs lost in females and the third leading cause in males^3^.

Large-artery atherosclerotic (LAA) stroke is the most prevalent subtype of ischemic stroke, notably in the Asian population, where it accounts for around 33%. Furthermore, LAA is the subtype with the highest yearly growth rate, at 5.7% ^4^. There are several risk factors that are related to LAA stroke occurrence. Traditional risk factors include age, sex, hypertension, diabetes mellitus, dyslipidemia, family history of cardiovascular diseases, current smoking, binge alcohol consumption, and obesity^5,6^. In addition, a number of recently emerging risk factors include the gut microbiome and gut microbiota-metabolites^5,7^. Emerging evidence suggests that gut microbiome, their metabolites and immune response play a crucial role in the pathogenesis of LAA stroke and may become a therapeutic target for its treatment^7,8^.

Gut microbiota, including bacteria, fungus and virus, are microorganisms that inhabit numerous areas of the body, primarily the gastrointestinal tract. The number of human microbiota is about 10^13^ - 10^14^ microbial cells. The proportion of microbial cells and human cells are around 1^9,10^. Firmicutes, Bacteroidetes, Actinobacteria, and Proteobacteria are the most prevalent bacterial phyla in the human gastrointestinal system; with approximately 90% being *Firmicutes* and *Bacteroides*. The *Firmicutes* include various genera such as *Lactobacillus and Ruminococcus. Bacteroidetes* are mainly represented by genera *Bacteroides* and *Prevotella*. The most abundant genus in phylum *Actinobacteria* are *Bifidobacterium*^11^.

The link of gut microbiota and stroke has not been previously elucidated. However, many researchers hypothesized that bacteria residing in the colon may be associated with stroke under the model of the Gut Microbiota-Brain Axis^12-14^. A previous cohort study examined how ischemic stroke altered the abundance of gut microbiota independent of age, typeII DM and hypertension^8^. In top-down signaling, Lactobacillus ruminis increased accordingly with markers of inflammation in stroke patients and SCFAs (Short-Chain Fatty Acids) were decreased. Furthermore, in animal model, occlusion of the MCA for 60 minutes increased intestinal permeability and produced gut dysbiosis^12,13^. However, the etiopathogenesis between gut microbiota and stroke is unclear as well as there are limit clinical studies focusing on this association. Therefore, the purpose of our study is to establish an association between the changes in gut microbiota and ischemic stroke patients, with a particular emphasis on LAA subtypes due to their high prevalence, in order to shed light on how gut microbiota may be a potential therapeutic target for ischemic stroke.

## Methods

A cross-sectional pilot study was conducted among 15 patients with LAA who were admitted at the stroke unit and 15 asymptomatic persons who presented at an outpatient clinic at the King Chulalongkorn Memorial hospital between year 2019 and 2020. At enrollment, participants were required to provide information on multiple lifestyle variables and medical history. Inclusion criteria were acute ischemic stroke patients aged over 45 years who were admitted to the Stroke Unit at King Chulalongkorn Memorial Hospital (KCMH), diagnosed as stroke caused by large artery atherosclerosis by neurologist using the TOAST classification system with symptom onset within 48 hours. Exclusion criteria were patients who received antibiotics within 1 month before stool collection, patients who had underlying diseases including metastatic cancer, autoimmune diseases, immunodeficiency, renal failure, chronic heart failure and Parkinson’s disease. For the control group, inclusion criteria were volunteers who were age and sex-matched with the stroke group. The exclusion criteria for the control group were the same as for the stroke group. Overall, 29 patients participated in the study, 14 in the stroke group and 15 in the control group. The study was approved by the Institutional Review Board of the Faculty of Medicine, Chulalongkorn University (IRB 029/62). All findings were reported in accordance with the STROBE guidelines (Strengthening the Reporting of Observational Studies in Epidemiology) for cross-sectional studies^15^.

### Baseline assessment

All participants underwent a comprehensive assessment that measured the followings: (1) demographic characteristics including age, sex, BMI; (2) potential risk factors for ischemic stroke, such as hypertension, dyslipidemia, diabetes mellitus, atrial fibrillation, ischemic heart disease, a smoking habit, a history of stroke, a family history of ischemic heart disease/ischemic stroke and alcohol consumption; (3) NIHSS score to assess severity of ischemic stroke (4) onset of acute ischemic stroke (5) laboratory variables, including lipid profile, HbA1C, creatinine, uric acid; (6) brain imaging and vascular imaging including computed tomography (CT) scan and computed tomography angiography (CTA); and (7) assessment of current medications (antibiotic, anti-hypertensive drugs, statins, proton pump inhibitors/H2 blockers, anti-thrombotic drugs

### Assessment of acute ischemic stroke

We categorized patients with acute ischemic stroke depending on TOAST classification which include Large-artery atherosclerosis (LAA), Cardioembolism, Small vessel occlusion, Stroke of other determined etiology and Stroke of undetermined etiology^16^. We only included LAA patients with clinical and brain imaging findings of either substantial (>50%) stenosis of a the major cerebral artery, presumedly related to atherosclerosis.^16^.

### Biochemical assays

Blood samples were obtained from patients with stroke at the time of admission and from control subjects during the outpatient clinic visit. Serum levels of glycated hemoglobin (HbA1c), high-density lipoprotein (HDL) cholesterol, low-density lipoprotein (LDL) cholesterol, triglyceride (TG), glucose, blood urea nitrogen, creatinine and uric acid were measured.

### Gut microbiome

Fecal samples of all stoke participants were collected within 1 days after admission and stored in –80 C within 60 minutes at the Thai Red Cross Emerging Infectious Disease Health Science Center laboratory by research assistants. Also, fecal samples from healthy individuals were collected at outpatient clinics and were sent by the same process. DNA extraction from stool samples were carried out, amplified and sequenced.

### Bacterial DNA extraction

Approximately 200 mg of stool samples were resuspended in 1 ml of InhibitEx buffer, incubated at 70°C for 5 minutes, and centrifuged at 20,000xg for 1 minute. The QIAamp fast DNA stool mini kit (QIAGEN) was used to extract bacterial DNA from 200 microliters of aqueous phase per the manufacturer’s instructions. All of the extracted DNA was quantified using the Qubit dsDNA HS test kit on a Qubit 4 Fluorometer (Life Technologies).

### 16S rRNA (V3-4) amplification and sequencing

The PCR products were separated on agarose gel electrophoresis, followed by gel extraction and purification using NucleoSpin gel and a PCR clean up kit (Macherey-Nagel). The indices of Illumina were applied to both ends of PCR products so that samples could be multiplexed. The indexed PCR products were then purified using Agencourt AMPure XP beads (Beckman Coulter, Inc.) and concentration was measured with a Qubit dsDNA HS test kit (Life Technologies). Prior to library pooling and sequencing, the accurate size of indexed libraries was determined through QIAxcel capillary electrophoresis (QIAGEN) utilizing paired-end (2×301 bp) sequencing on Illumina MiSeq with Illumina V3 reagent kit.

### Diversity Calculations

The sample-taxa frequency table was re-summarized at the phylum and genus levels by summing read counts belonging to the same phylum or genus together. Operational taxonomic units (OTUs) with unassigned phylum or genus were discarded from further analyses at the respective levels. Shannon alpha-diversity indices was calculated for each sample based on their mathematical definitions using vegan R package [27]. We used taxonomic data at the genus, family and order level to conduct a principle coordinates analysis based on Bray-Curtis dissimilarity using the Phyloseq R package. PERMANOVA (Adonis) was used to test for differences in Bray-Curtis beta diversity by stroke diagnosis. To evaluate beta-diversity structure, a phylogenetic tree containing all identified taxa was reconstructed using the phylogeny module in QIIME2.

### Statistical Analysis

Descriptive data analysis was performed using SPSS Statistics version 21 (SPSS Inc., Chicago, IL, USA). Categorical data were expressed as numbers and percentages. Normally distributed continuous data were presented as means with SD, while non-normally distributed continuous data were reported as medians with inter-quartiles range. A P value of less than 0.05 was considered as statistically significant.

We examined the association between each gut microbial genus and acute ischemic stroke using generalized linear mix effect model with multivariate adjustment for underlying disease including diabetes mellitus, hypertension and dyslipidemia, lifestyle variables including age, BMI, and alcohol intake. Analyses were performed using R version 4.0.1:

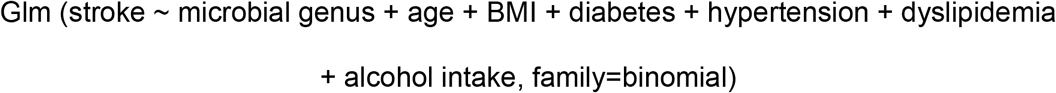

The circular phylogenetic tree was generated using GraPhlAn in Python version 3.6.4 to illustrate the relationship between each microbial species and stroke diagnosis.

## Results

There were 29 patients in the study. The average age of the stroke group being 61.1± 7.1 and that of the control group being 59.2± 8.2. Twelve out of 14 stroke patients (85.7%) and 12 out of 15 control group (80%) were male. Hypertension and dyslipidemia were the most prevalent traditional risk factors for ischemic stroke in both groups, accounting for 18 out of 29 participants (62%). Baseline characteristics of the patients were shown in **Table 1**.

**Table 1.** Baseline characteristics of included participants

| Characteristic | Stroke patients<br>N= 14 | Control<br>N=15 | P-value |
| --- | --- | --- | --- |
| Male: n (%) | 12 (85.7) | 12 (80) | 1.000 |
| Age (mean±SD) | 61.1± 7.1 | 59.2± 8.2 | 0.720 |
| Risk factors: n (%) |  |  |  |
| History of diabetes mellitus | 1 (7.1) | 2 (13.3) | 0.543 |
| History of hypertension | 5 (35.7) | 4 (26.7) | 0.690 |
| History of dyslipidemia | 4 (28.6) | 5 (33.3) | 0.690 |
| Smoking | 2 (14.3) | 0 (0) | 0.143 |
| Previous stroke | 1 (7.1) | 0 (0) | 0.390 |
| Lab investigation (mean±SD) |  |  |  |
| Total cholesterol | 181.9±42.9 | 179.3±28.2 | 0.842 |
| HDL | 38.1±10.4 | 46.1±17.2 | 0.145 |
| LDL | 115.9±40.1 | 109.4±25.5 | 0.948 |
| TG | 139.9±38.6 | 119.0±33.2 | 0.941 |
| HbA1c | 5.7±0.3 | 5.9±0.7 | 0.416 |
| NIHSS on admission (mean±SD) | 1.28±1.09 | - | - |

The majority of intestinal bacteria discovered in both patient and control groups were Clostridia class, which belong to the phylum Firmicutes, according to taxonomic classification at the class level **(Figure 1)**. A significant difference in Bacteroidia was found in approximately 7% of the stroke group but only 2% of the control group (Mann Whitney U test; P 0.014). **(Supplementary figure 1)**.

**Figure 1:**
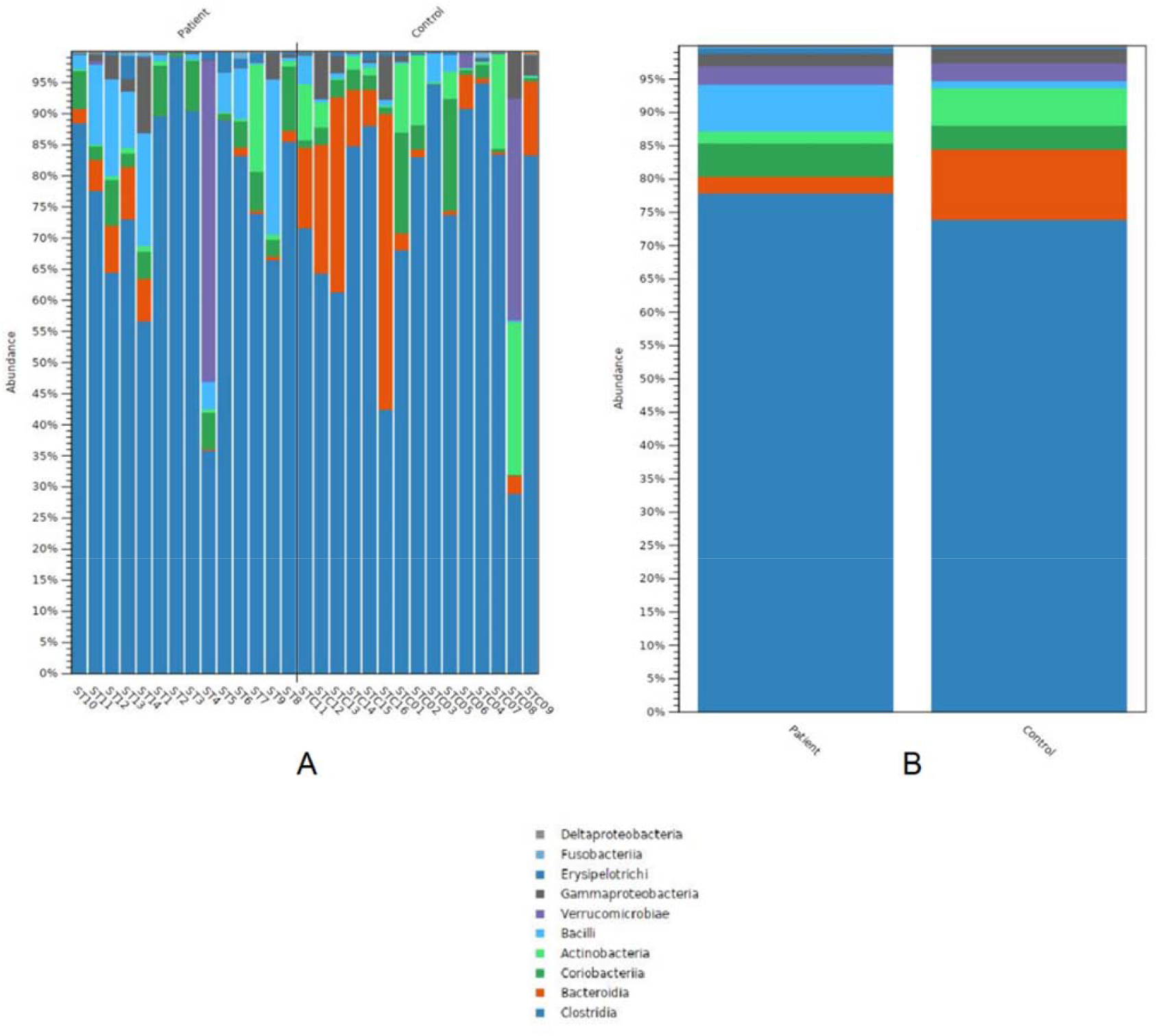
Class-level taxonomic overview of the gut microbiota. (A) Summary of gut microbiota in each specimen (B) Summary of gut microbiota between stroke and control group

Stool samples were underwent 16S rRNA sequencing and analyses. We found no significant differences between stroke and control groups in alpha-diversity index (Shannon) (P-value 0.852) **(Supplementary figure 2)**. Beta-diversity evaluation using PCoA showed significant differences at the order family and genus levels between strokes and controls (P-value 0.017, P-value 0.011 and P-value 0.003, respectively) **(Figure 2)**.

**Figure 2:**
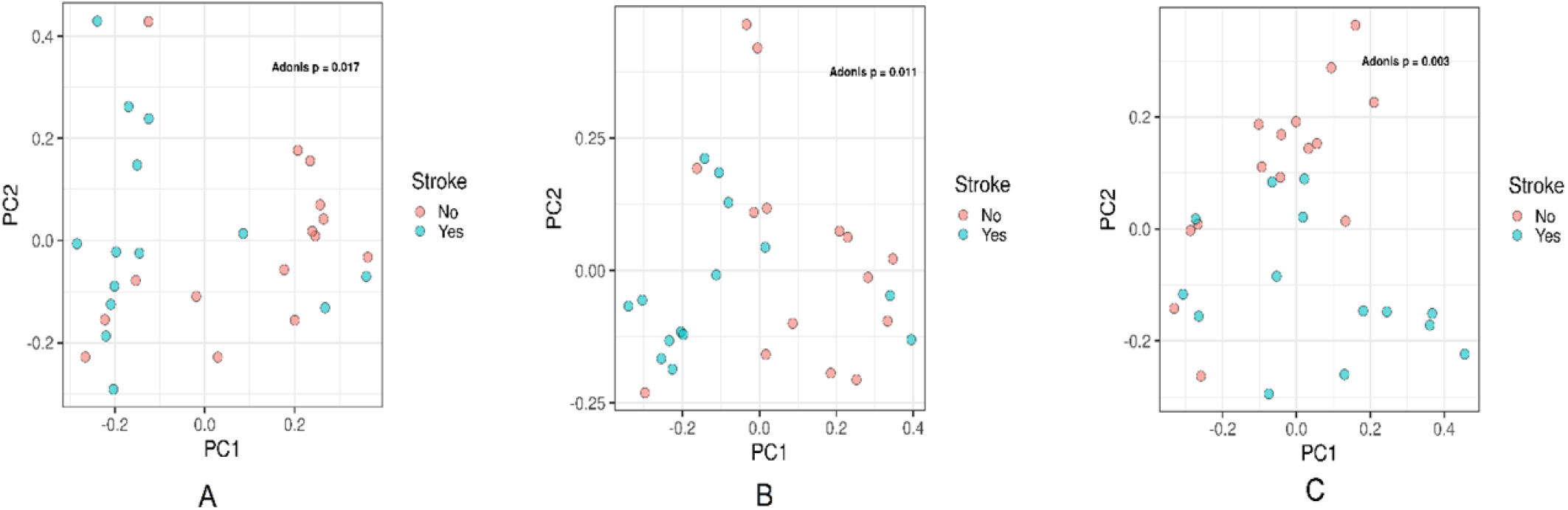
Beta-diversity of bacterial order (A), family (B), and genus (C) in stroke and control group. PCoA based on Bray–Curtis dissimilarity was performed with the significant differences in gut microbial community structure at the order (P-value 0.017), family (P-value 0.011) and genus levels (P-value 0.003) between stroke and control group.

Using generalized linear mix effect models, we found that 6 genera were significantly associated with stroke after multivariate adjustment (P-value <0.05) (4 positive association and 2 negative association). *Ruminococcus spp*. (Beta 18.70, P-value 0.017), *Streptococcus spp*.(Beta 9.25, P-value 0.019) and *Actinomyces spp*.(Beta 63.37, P-value 0.02) And *Dorea spp*.(Beta 46.10, P-value 0.021) showed positive association while *Bifidobacterium spp*. (Beta -4.4, P-value 0.04) and *Faecalibacterium spp*. (Beta -7.16, P-value 0.041) showed negatively association with stroke **(Figure 3)**.

**Figure 3:**
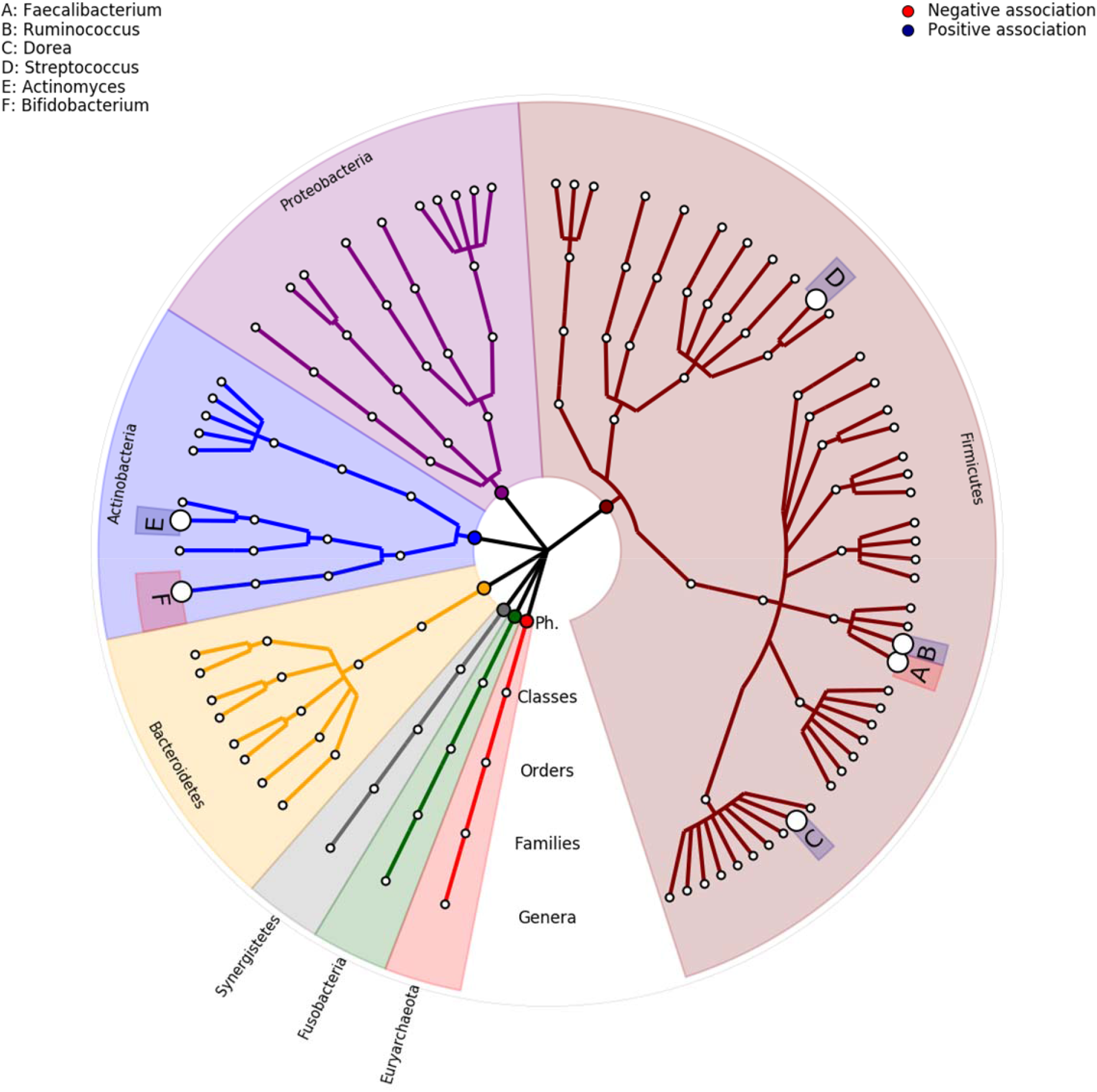
GraPhlAn visualization of the annotated phylogenies and taxonomies compared between stroke and control group.

## Discussion

In this study, a correlation was found between large-artery atherosclerotic stroke and gut microbiome diversity and as well as specific microbial genus. Bray– Curtis dissimilarity was decreased in gut microbial community structures at the order, family and genus in stroke group. Although the underlying mechanism remains unclear, it was hypothesized that communication from gut-microbiota to acute ischemic stroke or from acute ischemic stroke to gut-microbiota or both may be involved^12,13^. Regarding gut-microbiota to stroke communication, low-diversity dysbiosis may modify the metabolic flow of bacteria and their direct interactions with the host immune system^17^. Due to low microbial diversity, many anaerobes in a healthy gut convert complex carbohydrates to short chain fatty acids (SCFAs)^17^.

Owing to the anti-inflammatory and cholesterol-blocking effects of SCFAs^18^, patients with reduced microbial diversity are more susceptible to LAA stroke than the control group. Regarding the latter hypothesis that stroke leads to gut microbial dysbiosis, a recent study on mice revealed that ischemic stroke affects the gut microbiome, reduces microbial diversity, and boosts the immune system ^19^. Also, In an another mice study, reduced microbiome diversity was also found as a characteristic of post-stroke dysbiosis, which was related with decreased intestinal motility^8^. This finding was supported by some hypotheses, the first of which is that the autonomic nerve system mediates the effect of stroke on dysbiosis. Houlden et al. determined that stroke affected the composition of cecal microbiota which these microbiome changes were mediated by the release of noradrenaline from the autonomic nervous system, affecting the synthesis of cecal mucoproteins and the quantity of goblet cells^20^. In addition, it was hypothesized that the post-stroke stress response increases intestinal permeability via the production of corticotropin-releasing and glucocorticoid hormones, resulting in enhanced bacterial translocation in the gut^21,22^.

We found a negative association between *Bifidobacterium* spp. and stroke. *Bifidobacterium spp*. has been commonly referred to as beneficial bacteria that perform necessary functions in the human colon^23^ and was developed as a widely used probiotics. ^24^. Nowadays, *Bifidobacterium spp*. probiotics are now commonly used to treat irritable bowel syndrome and ulcerative colitis by altering the composition of the microbiota in the gut. ^25^. Decreased numbers of these species in the colon have been linked to numerous diseases, such as antibiotic-associated diarrhea, irritable bowel syndrome, inflammatory bowel disease, obesity, allergies, and regressive autism. *Bifidobacteria spp*. has served multiple purposes, including the development of the immune system in early life, maintenance of the intestinal barrier, and protection against pathogens^26^.

Regarding *Bifidobacterium spp*. and stroke, a number of research have demonstrated that *Bifidobacterium* treatment can effectively enhance the long-term rehabilitation of mice with cerebral ischemia^27^. It was hypothesized, while the mechanism remained unknown, that *Bifidobacterium spp*. generated metabolites of short-chain fatty acid (SCFA), which can decrease inflammation and improve stroke recovery^28,29^. Also, *Bifidobacterium*-treated mice exhibited an upsurge in a range of metabolites, including prostaglandin B1, which may promote stroke recovery^30,31^.

*Faecalibacterium spp*. abundance were lower in the stroke group than in the control group, according to our findings. *Faecalibacterium prausnitzii* is one of the most essential gut microbiota components in the human colon, which has been considered a bioindicator of human health^32^. Changes in the abundance of *F. prausnitzii* have been linked to dysbiosis in a variety of human disorders. ^33-35^. As a Butyrate-producing bacteria, it was found to be reduced in cardiovascular disease and metabolic syndrome. Butyrate is an essential fatty acid with a short chain that promotes intestinal health^36^. In addition, this microbe possesses a variety of anti-inflammatory and metabolic properties, which give it an important role in human health^37^. Butyrate protects the intestinal lining, thereby preventing infections from entering the body via the gastrointestinal tract. It stimulates the growth of villi and the production of mucin, a protective gel that coats the digestive tract lining^38^. Concerning stroke, studies on mice have demonstrated a correlation. In a previous mice study from China, for instance, *Faecalibacterium* was used for transplantation. ^39^. These bacteria decreased post-stroke neurological deficits and inflammation and increased SCFA concentrations in the stomach, brain, and plasma of aged mice with stroke. ^39^. The majority of studies have shown an association between *F. prausnitzii* and stroke; however, our study only analyzed genus data but not species data due to 16S NGS analysis. Therefore, future studies should conduct 16S metagenomic analysis in order to determine the link between *F. prausnitzii* and stroke.

Our findings indicated that four microbial genera (*Ruminococcus spp*., *Streptococcus spp*., *Actinomyces spp*., *and Dorea spp*.) were positively correlated with stroke group. The relationship between these microbial genera and stroke has been demonstrated in a small number of studies. *Ruminococcus spp*. And *Streptococcus spp*. have been associated with metabolic syndrome and cardiovascular disease in certain research. First, Kurilshikov et al. discovered the association between the *Ruminococcus* species and the onset of cardiovascular diseases^40^. The production of L-methionine by *Ruminococcus* species is associated with cardiovascular characteristics in obese people^41^. Second, *Streptococcus spp*., a morbid oral bacterium species, has also been shown to be raised in hypertension ^42^, and atherosclerotic cardiovascular disease ^*43,44*^. Finally, *Actinomyces spp*. was discovered to be prevalent in obese adolescents^45,46^, and a positive association was also identified between *Dorea spp*. and BMI and blood lipids^40^. Consequently, these four genera may have an impact on LAA stroke via the cardiovascular disease pathway, thereby contributing to the development of stroke. Even while no studies showed the potential mechanism of these microbial genera, future research should investigate the relationship between these microbial genera and large-artery atherosclerotic stroke.

Our study had some limitations. First, our study had small sample size which may affect our results. Second, we have no metabolomic data in our databases and the causality between this association have not been conducted due to the cross-sectional design of the study. For future research to clarify the relationship between gut microbiota, its metabolites, and stroke, cohort studies with a metabolomic profile should be implemented. Finally, our research is based on the results of 16S rRNA sequencing within the limitations of resolution. A subsequent study is planned to compare these findings, which should broaden our understanding through functional microbiome analysis. The long-term objective of this research is to develop metatranscriptomic and metatranscriptomic analysis between gut microbiome and large-artery atherosclerotic stroke.

## Conclusion

Our findings suggests that patients with large-artery atherosclerotic stroke had a decreased beta-microbiome diversity, and certain gut microbiota genera may be related to large-artery atherosclerotic stroke. Future implications of this study could include the development of targeted interventions to modulate gut microbiota in order to improve outcomes for patients with large-artery atherosclerotic stroke.

## Data Availability

All data generated or analysed during this study are included in this article. Further enquiries can be directed to the corresponding author.

## Abbrevations

BMI: Body mass index
CT: Computed tomography
CTA: Computed tomography angiography
LAA: Large-artery atherosclerosis
NIHSS: National Institute of Health Stroke Scale
PCR: Polymerase Chain Reaction
SD: Standard deviation
TOAST: Trial of Org 10,172 in Acute Stroke Treatment

## Statements

The study was approved by the Institutional Review Board of the Faculty of Medicine, Chulalongkorn University (IRB 029/62).

## Statement of Ethics

All participants provided written informed consent at the time of data collection. The study was approved by the Institutional Review Board of the Faculty of Medicine, Chulalongkorn University (IRB 029/62) and adheres to the tenets of the Declaration of Helsinki. All participants’ data were fully anonymized.

## Author Contributions

Chatpol Samuthpongtorn: data gathering, interpretation of data, draft writing, editing, and revision of manuscript.

Abhinbhen W. Saraya: study conception, supervision, editing, and revision of manuscript.

Yutthana Joyjinda: interpretation of data

Apaporn Rodpan: interpretation of data

Nijasri C. Suwanwela: study conception, interpretation of data, supervision, editing, and revision of manuscript.

## Acknowledgments

None

## Supplemental Material

Figures S1–S2

